# Spatially regulated designs of incidence surveys can match the precision of classical survey designs whilst requiring smaller sample sizes: the case of snakebite in Sri Lanka

**DOI:** 10.1101/2022.04.24.22274231

**Authors:** Dileepa Senajith Ediriweera, Tiloka de Silva, Anuradhani Kasturiratne, Hithanadura Janaka de Silva, Peter John Diggle

## Abstract

**Background:** Snakebite envenoming is a neglected tropical disease. Data from the worst affected countries are limited because conducting epidemiological surveys is challenging. We assessed the utility of inhibitory geostatistical design with close pairs (ICP) to estimate snakebite incidence.

**Methods:** The National Snakebite Survey (NSS) in Sri Lanka adopted a multistage cluster sampling design, based on population distribution, targeting 1% of the country’s population. Using a simulation-based study, we assessed predictive efficiency of ICP against a classical survey design at different fractions of the original sample size of the NSS. We also assessed travel distance, time taken to complete the survey, and sensitivity and specificity for detecting high-risk areas for snake envenoming, when using these methods.

**Results:** A classical survey design with 33% of the original NSS sample size was able to yield a similar predictive efficiency. ICP yielded the same at 25% of the NSS sample size, a 25% reduction in sample size compared to a classical survey design. ICP showed >80% sensitivity and specificity for detecting high-risk areas of envenoming when the sampling fraction was ≥20%. When ICP was adopted with 25% of the original NSS sample size, travel distance was reduced by >40% and time to conduct the survey was reduced by >75%.

**Conclusions:** This study showed that snakebite envenoming incidence can be estimated by adopting an ICP design with similar precision at a lower sample size than a classical design. This would substantially save resources and time taken to conduct epidemiological surveys and may be suited for low resource settings.

**Key messages:**

- Inhibitory geostatistical design with close pairs (ICP) for incidence surveys can match the precision of classical survey designs at lower sample sizes.
- The ICP design showed a lower predictive variance than classical design, indicating ICP designs were able to produce more reliable predictions.
- The ICP design showed a lower time to complete the survey than the classical sampling method.
- Although primary sampling units in the ICP design maintain a minimum distance between two units, ICP did not increase travel distance compared to the classical survey design.
- Resource requirements and time to complete surveys can be reduced without increasing the distance to travel by adopting ICP design for epidemiological surveys.

## Introduction

Snakebite envenoming is a neglected tropical disease, mainly affecting poor rural communities ^1^. Robust survey data on snakebite incidence are scarce in most of these countries, and hospital records are known to underestimate the burden ^2,3^. Only a few community-based surveys have been conducted to estimate snakebite incidence at national level in these countries due to the lack of resources and logistical difficulties ^3^.

Sri Lanka is a tropical island nation with a high incidence of snakebite envenoming ^4^. The island is located in the Indian Ocean, with a land area of 65 000 km^2^ and a population exceeding 20 million. There are 100 terrestrial snake species in the country ^5^. Sri Lanka was the first country to estimate the country-wide community incidence of snakebite and envenoming by conducting a national snakebite survey (NSS) in 2012-13 ^3^. The NSS adopted a multistage cluster sampling design, targeting 1% of the total population with cluster selection based on the population distribution; more clusters were chosen in population-dense areas and vice versa.

From a spatial prediction point of view, the selection of clusters in NSS could be inefficient due to the risk of oversampling and under-sampling in areas with high and low population densities, respectively ^6^. Therefore, the spatial predictions are likely to be associated with varying degrees of precision over the region of interest, with especially low precision in areas with low population densities ^7^. This can be addressed by adopting geostatistical sampling methods such as inhibitory close pair (ICP) designs. In ICP, primary sample units are separated by a given minimum distance, which safeguards against the geographically unbalanced selection of sampling units as in the NSS ^6^.

The NSS in Sri Lanka was feasible given the country’s relatively small geographical size. Even so, the survey took 11 months to complete and was logistically challenging. As such, undertaking national surveys on the same scale in other countries with high snakebite burdens and resource constraints (or even repeating the same survey within Sri Lanka) would be very challenging. Therefore, this study aimed to apply a geostatistical sampling method, specifically ICP, to estimate the incidence of snakebite envenoming and highlight the utility of geostatistical designs to achieve a similar precision to more resource intensive sampling methods such as those based on population distribution.

## Method

### Sampling

The national snakebite survey (NSS) in Sri Lanka is one of the largest surveys done in neglected tropical disease settings. Therefore, we considered NSS for this simulation study. The NSS was a country-wide community-based cross-sectional survey conducted between August 2012 and June 2013. It was the first survey to be conducted at a national level to estimate snakebite incidence. Given that the true snakebite incidence in Sri Lanka was unknown at the time, approximately 1% of the population of the country was sampled.

#### Original sampling design adopted in NSS : Classical survey design

Sri Lanka has nine provinces and 25 districts. The last census of population and housing carried out before the NSS was conducted in 2001 and covered only 18 districts, where the population was found to be 18.8 million living in 3.9 million households. Based on these numbers, the investigators estimated the population of 2012 to be 20 million and the number of households to be 4.5 million. Accordingly, to cover 1% of the population, 45 000 households were sampled, divided equally among the nine provinces.

The National Snakebite Survey adopted a classical survey design (i.e. multistage cluster sampling method) to recruit participants. The *Grama Niladhari* (GN) divisions are the smallest administrative unit in the country; there are 14022 such divisions in the country. The GN divisions were considered as the first-stage sampling units (e.g. clusters), with 125 GN divisions selected from each province. The 125 GN divisions were then proportionately allocated to the districts within a province based on the population distribution between districts (i.e. proportional allocation to strata). The GN divisions within a district were selected by simple random sampling. The sampled GN divisions are shown in Figure 1.

**Figure 1:**
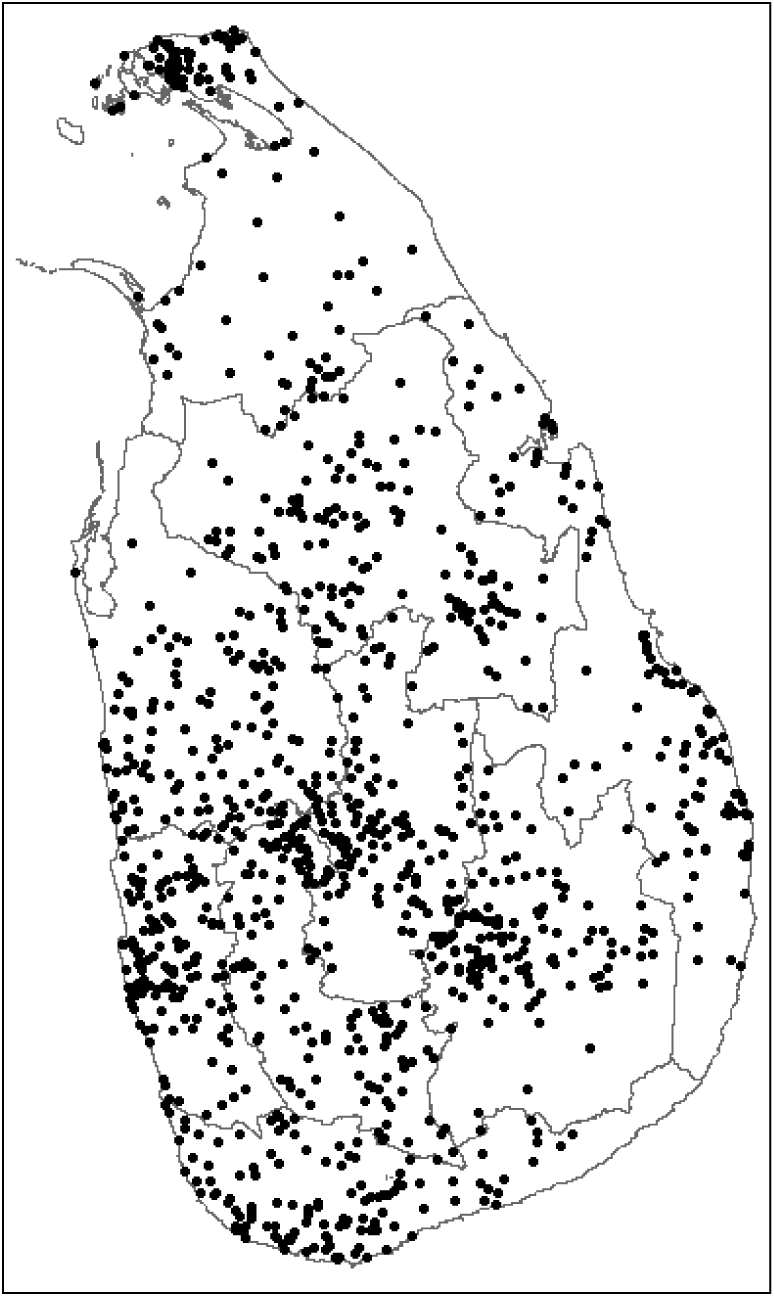
The sampled *Grama Niladhari* divisions (i.e. clusters) in the National Snakebite Survey. Grey lines demarcate the nine provinces in Sri Lanka.

A household was considered as the second-stage sampling unit, with 40 households sampled from each GN division. The first household was randomly selected from the electoral register, after which proximity selection was adopted to select the next 39 households (i.e. “next nearest”). All members of the household were recruited for the survey, which collected data on all snakebites as well as snakebites with significant envenoming (presence of local tissue necrosis at the site of the bite, presence of neurotoxicity or bleeding manifestations) that took place within the previous 12 months. Details of the survey have been published elsewhere ^3^.

#### Inhibitory geostatistical design with close pairs (ICP)

Snakebite incidence estimated using data from the NSS showed substantial spatial variation. The previous geostatistical models indicated the existence of a spatial correlation between events and a presence of a non-negligible nugget effect in the residual covariance structure. Since an inhibitory geostatistical design alone cannot reliably estimate the covariance structure associated with a combination of spatial and non-spatial random effects, we chose an inhibitory geostatistical design with close pairs (ICP) to select the first-stage sampling units (i.e. GN divisions) in this study ^6^. ICP has two types of sample points; primary sample points which maintain a minimum distance between any given two locations and supplementary sample points (i.e., close pairs) which are chosen within a given radius of primary sample points.

The centroids of all the 14022 GN divisions in the country formed our sampling frame. The steps followed in the ICP sampling method are outlined below, where *n* is the total number of required sample points, *δ* is the minimum distance between any given two primary sample points, *k* is the number of supplementary sample points (i.e. close pairs), and *ζ* is the radius of the disc where a supplementary or paired point is added.

1. Draw a sample of locations *x*_*i*_: *i = 1,…*.,*n-k* randomly from the sampling frame (i.e.14022 GN divisions)
2. Set *i* =1;
3. Calculate the minimum distance, *d*_*min*_, from *x*_*i*_ to all other *x*_*j*_ in the sample
4. If *d*_*min*_ >= *δ*, increase *i* by 1 and return to step 3 if *i* <= *n*, otherwise stop.
5. If *d*_*min*_ < *δ*, replace *x*_*i*_ with a new location drawn at random from the sampling frame and return to step 3
6. Sample *k* locations from *x*_*1*_,*…*..*x*_*n-k*_ without replacement and denote them *x*_*j*_^*\**^, *j* = *1,…*.,*k;*
7. For, *j* = *1,…*.*k*; x_n-k+j_ is chosen at random from all as-yet unsampled locations within the disc with centre x_j_^*^ and radius *ζ*.

In our study, k was selected as 20% of the total number of required sample points, *n, δ* was set at the maximum value for which it was possible to sample *n-k* locations, and *ζ* was selected as one third of *δ*.

### Simulation study

We undertook a simulation study to assess the predictive efficiency of classical survey design against ICP with different fractions (i.e. thinning) of the original NSS sample size. We followed the same modelling approach to estimate the snakebite envenoming incidence using the same explanatory variables as in the original study ^3^.

The true bite incidence in Sri Lanka is unknown, therefore, predictive incidence maps which were already published based on the original NSS data were assumed to be the gold standard. The Medical Officer of Health (MOH) divisions are the smallest administrative divisions in the public healthcare system in Sri Lanka. Public health initiatives are implemented through these divisions. We defined the MOH areas having an average bite incidence of more than the national incidence (i.e. 151 per 100 000) as high-risk MOH.

Incidence maps were then developed and predictive variances were calculated as follows;

1. Simulate the number of bites in the chosen GN divisions
2. Fit geostatistical models for the simulated bites in step 1 and develop predictive maps using a prediction grid surface
3. Calculate the average predictive variance (APV) over the prediction grid surface, sensitivity and specificity of detecting high-risk MOH areas of envenomation.

#### Geostatistical model

We used the same geostatistical model that was developed using the original NSS data ^3^ to model the simulated bites. At location *x*_*i*_, the number of bites, *y*_*i*,_ out of n_i_ individuals sampled are independent binomial events conditional on an unobserved spatial stochastic process *S(x)*, with probabilities *p(x*_*i*_), where

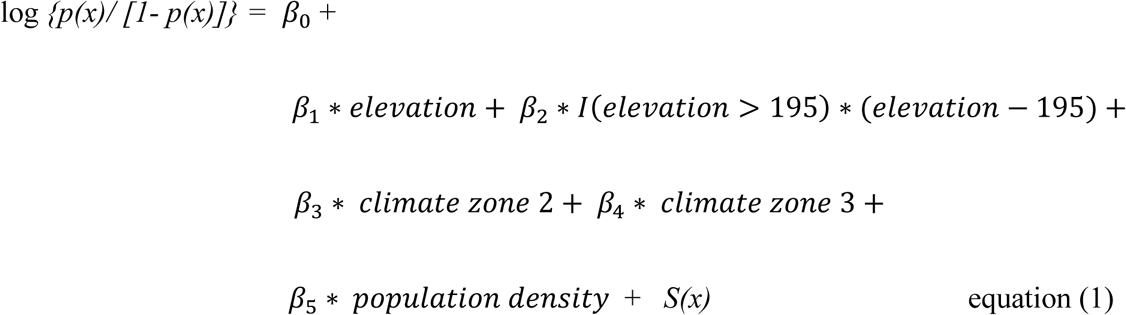

In equation (1), *I*(*elevation*> 195) is equal to one of elevation is greater than 195metres, and zero otherwise. The term *S(x)* captures any residual spatial variation after adjusting for the covariates and is modelled as a Gaussian process with mean zero, variance σ^2^ and correlation structure Corr Corr*[S(x), S(x’)]*= exp *(-u/φ)*,, where *u* is the distance between *x* and *x’*, and *φ* represents the scale of spatial correlation. Hence, the conditional mean number of bite incidence at location *x*_*i*_ depends on the explanatory variables observed at location *x*_*i*_ and on *S(x*_*i*_), and *p(x)* is the probability that a person at location *x* will experience a bite ^8^.

#### Simulating the number of bites

Sri Lanka has 140 22 GN divisions. We selected different fractions of the original NSS sample size (i.e. 50%, 33%, 25%, 20% and 10%) to conduct the simulations for both classical survey design and ICP.

The simulation of bites was done as follows;

1. Draw a sample of GN divisions *c*_*i;*_ *i = 563 (50%), 372 (33%), 282 (25%), 225 (20%), 169 (15%), 113 (10%)* using each of the two sampling methods.
2. Extract bite incidence in the selected GN divisions from the gold standard incidence map
3. Draw a random number of bites (i.e. observations) at each selected location using the binomial distribution for 150 trials and bite probability extracted from step 2. Note that the NSS surveyed 40 households (i.e. second-stage sampling units) in each GN division which included approximately 150 people from a GN division (i.e. 3 – 4 people per household).
4. Extract the values of explanatory variables in equation 1 for the selected GN divisions from the respective raster maps.

#### Geostatistical modelling and predictive mapping

The simulated bites were modelled using binomial geostatistical models (equation 1). Parameter estimates of the models were obtained by the Monte Carlo maximum likelihood method ^8^. A predictive grid surface of 2 km by 2 km was developed for the entire country and plug-in spatial predictions were obtained for mapping the results. PrevMap package version 1.5.4 in R programming language was used to fit the geostatistical models ^9^.

#### Calculating average predictive variance, sensitivity and specificity

Both the original predictive map (i.e. the gold standard) based on the NSS and simulated maps used the same 2 km by 2 km predictive grid surface. The original predictive map and simulated maps were compared at each grid point and the average predictive variance (APV) was calculated as follows;

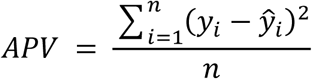

At location x_i_, *i* = 1,*2,…. n*, y_i_ is the actual bite incidence and *ŷ*_*i*_ is the bite incidence predicted by the simulations.

Subsequently, we calculated the sensitivity and specificity of detecting high-risk MOH divisions for each simulated map.

We replicated the above simulation 100 times for the classical survey design and ICP for each fraction of the original NSS sample size (i.e. 100%, 50%, 33%, 25%, 20% and 10%) to obtain the mean and standard deviation for APV, and the sensitivity and specificity of detecting MOH areas with high and low bite incidences.

### Distance calculation

The data collection of the NSS was carried out at the provincial level, with a single group completing data collection in one province before moving on to the other provinces. Because the ICP design imposes a minimum distance between sampling units, the travel distances involved for data collection within a province are unlikely to reduce in the same proportion as the reduction in sample size. Therefore, we estimated the travel distance for data collection within each province with ICP designs and compared it with the sampling strategy used in the NSS.

The distance travelled within a province was computed by randomly selecting a sampled GN division (through ICP or the original sampling strategy used in the NSS) from a given province and then using the *travelling salesman algorithm* in the TSP package on R programming language ^10^. This algorithm estimates the shortest route that covers all the selected GN divisions without any repetitions within a province. The sample selection and travel distance calculation procedures were then repeated 100 times to obtain mean travel distances for each ICP design from a random starting point.

The above computation requires a matrix of distances between each pair of GN divisions within a province. Given the cost constraints involved in estimating actual road distances for high-dimensional matrices, the distances were first approximated as the Euclidean (straight-line) distances between the centroids of a selected pair of clusters and then adjusted to better reflect road distances based on a district-level correction factor. The adjustment was based on the actual road distances from a randomly selected GN division to each of the remaining GN divisions within a district. This was more parsimonious than computing road distances between every pair of GN divisions within a district. For instance, in a district with 500 GN divisions, this approach required only 500 distances to be computed rather than 500^2^). The correction factor for a district was calculated as the average of the ratios between the road distance and Euclidean distance from the randomly selected GN division to the remaining GN divisions within the district. The actual road distances used for the calculation were obtained using the Distance Matrix application programming interface (API).

### Survey time calculation

In the NSS, the time taken to complete the survey in a household depended on whether a member of that household had been a victim of a snakebite or not. In households that did not have victims of snakebite, the average time taken to complete the survey was 10 minutes compared to an average of 40 minutes in households that did. Using these assumptions, we calculated the time taken to complete the survey for the simulated bites as described above. We replicated the above simulation 100 times for the classical survey design and ICP for each fraction of the original NSS sample size (i.e. 100%, 50%, 33%, 25%, 20% and 10%) to obtain the mean and standard deviation for time taken to complete the survey.

## Results

Figures 2 and 3 respectively demonstrate an instance of a sampled locations from classical survey design and ICP at different sampling fractions.

**Figure 2:**
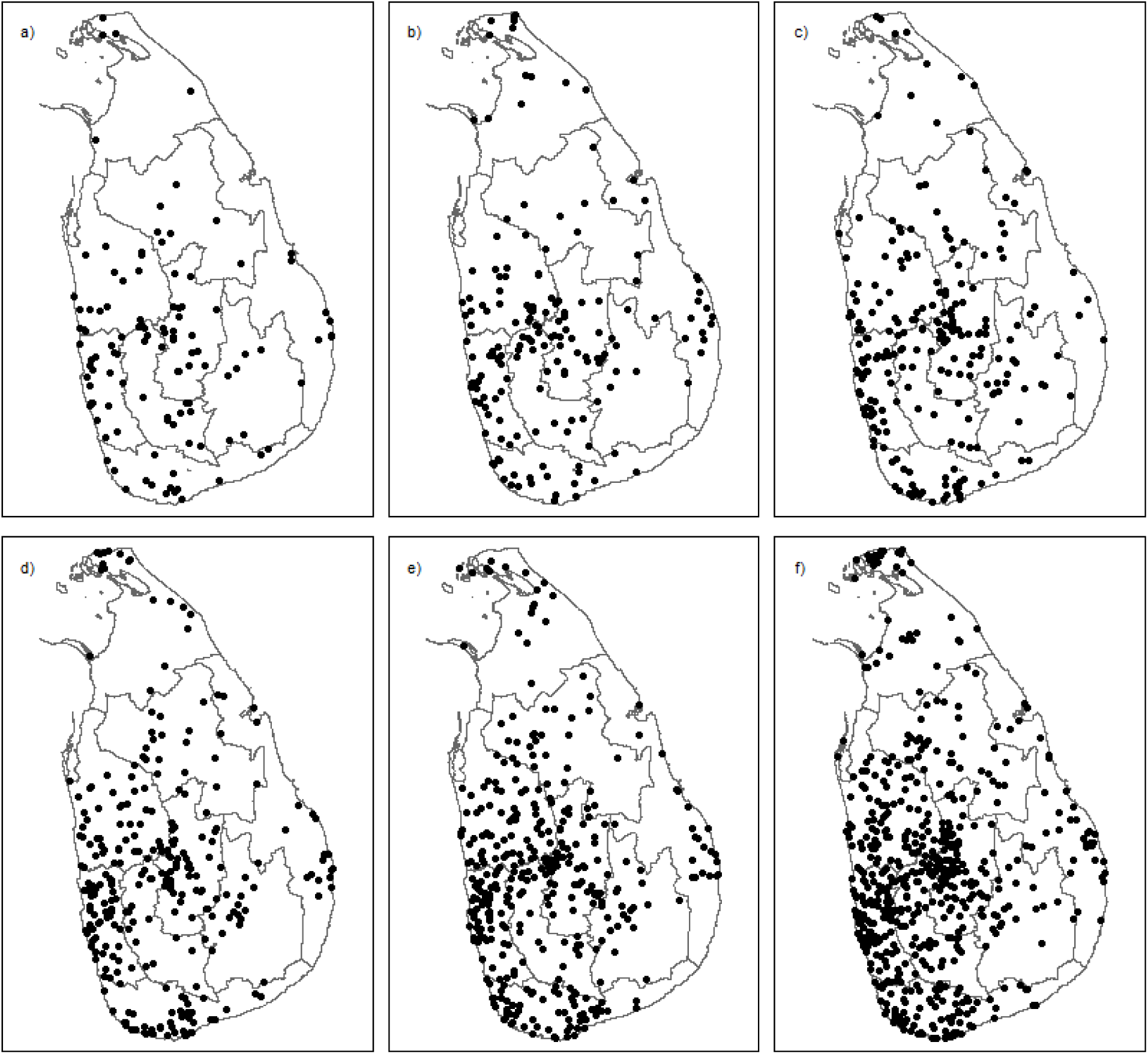
*Grama Niladhari* divisions selected by classical design. Panel a) 10%, b) 15%, c) 20%,d) 25%, e) 33% and f) 50% of the original sample size in the National Snakebite Survey.

**Figure 3:**
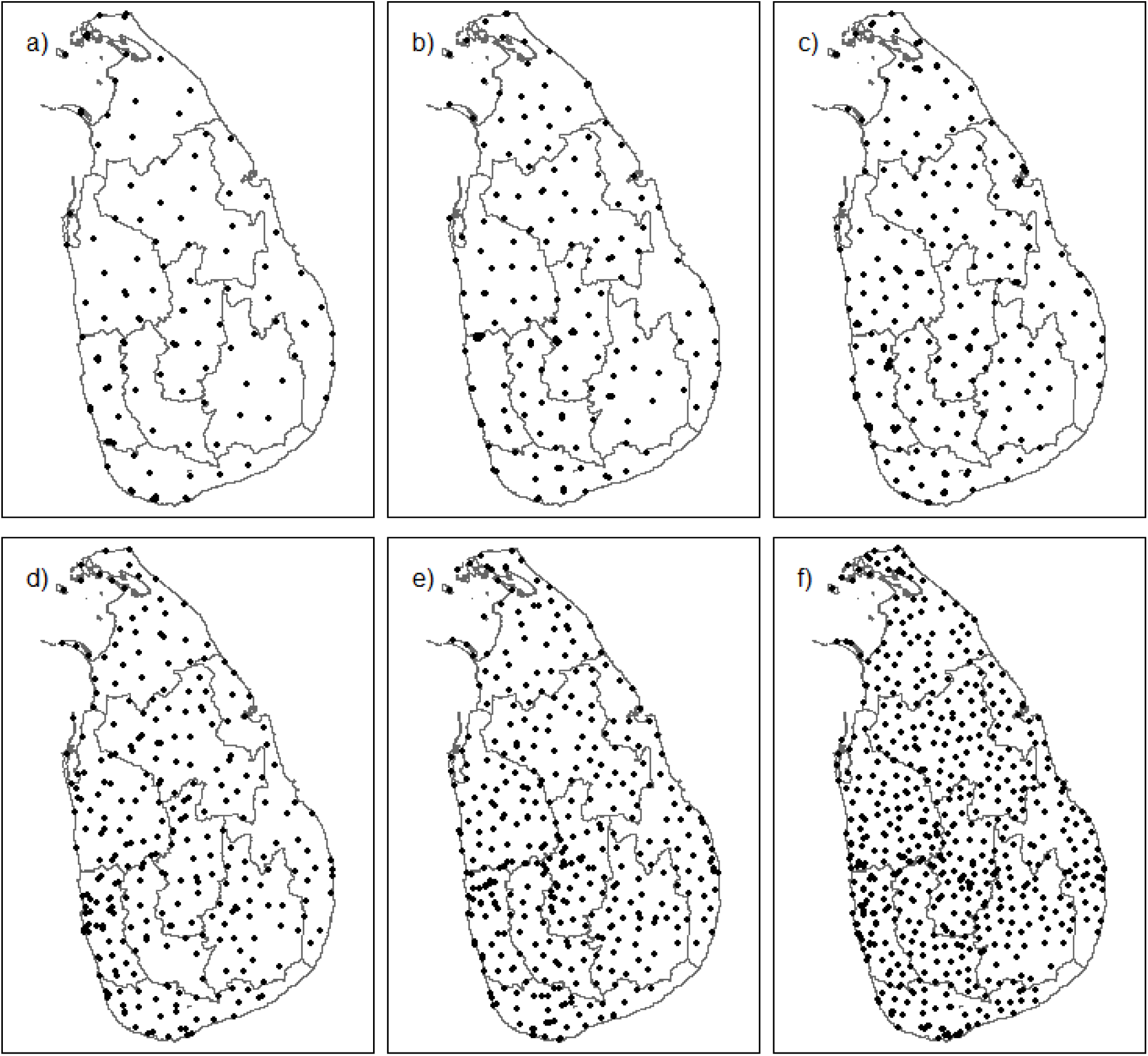
*Grama Niladhari* divisions selected by inhibitory geostatistical design with close pairs (ICP). Panel a) 10%, b) 15%, c) 20%, d) 25%, e) 33% and f) 50% of the original sample size in the National Snakebite Survey.

### Prediction variance

The APV of the 100 replicates of simulations are shown in figure 4. The APV of classical survey designs showed relatively higher deviation from the original sample size compared to ICP. The 95% confidence limits of the original sample size overlapped with the APV of 33% and 50% sampling fractions in classical survey designs. It overlapped with the AVP of 25%, 33% and 50% sampling fractions in ICP. This indicated that a similar predictive efficiency can be achieved with a sample fraction of 33% in a classical survey design and 25% in an ICP, which in turn represents a 25% reduction in the required sample size for an ICP design by comparison with a classical survey design. For all the sampling fractions considered, the 95% confidence limits of the variance ratio between classical design and ICP were substantially bigger, indicating the gains in precision achieved by ICP is better than by the classical designs.

**Figure 4:**
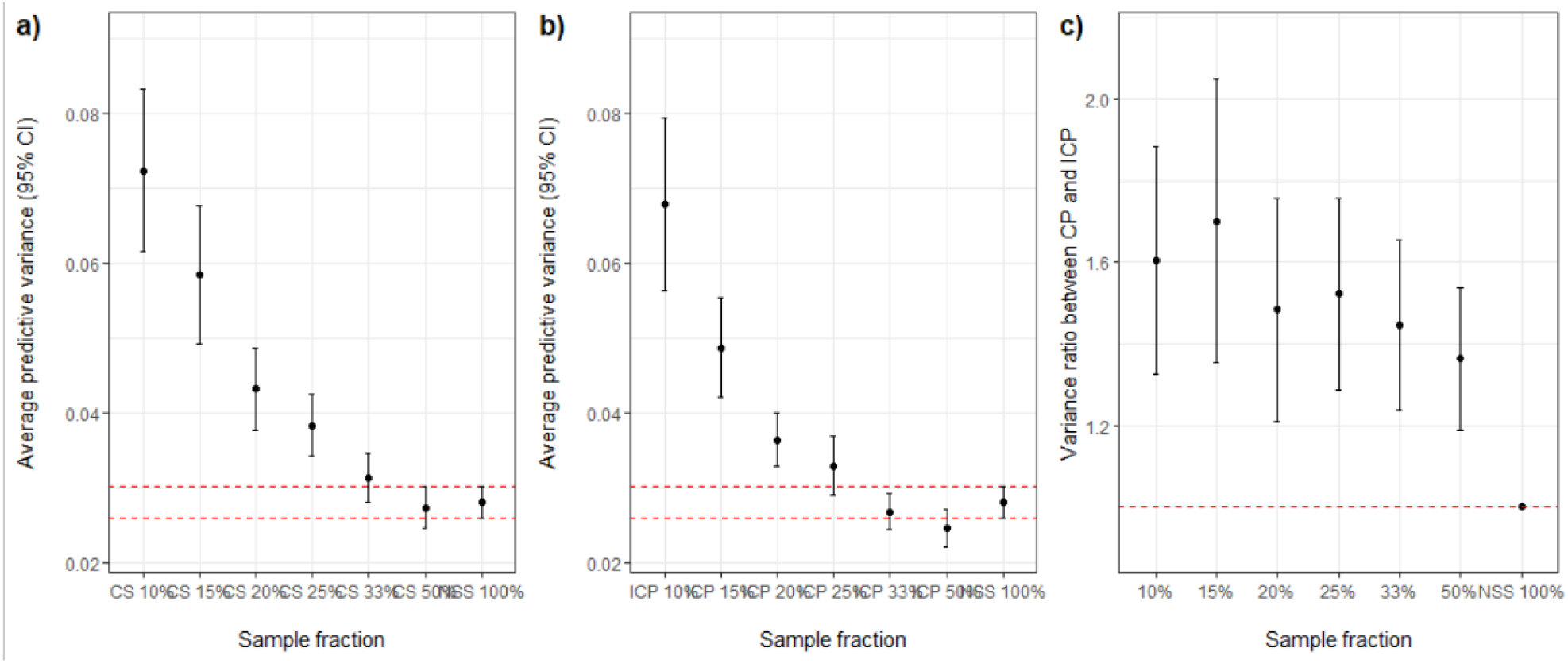
Results of the simulation. The average predictive variance (APV) of 100 replications of simulations a) classical design, b) inhibitory close pairs (ICP) design and c) variance ratio between classical design and ICP design with at 10%, 15%, 20%, 25%, 33%, 50% and 100% of the original sample size in the National Snakebite Survey (NSS). Red dashed lines indicate 95% CI of the APV of the NSS sampling in panel a) and b), and variance ratio of one in panel c).

### Sensitivity and specificity

Table 1 shows the sensitivity and specificity of the 100 replicates of simulations. The original NSS sampling showed 81.1% (95% CI: 79.5 – 82.6) sensitivity and 91.5% (95% CI: 89.9% – 93.0%) specificity in detecting high-risk MOH areas for snakebite envenoming. Classical survey designs showed a lower sensitivity and a higher specificity than ICPs with the same sample size. All the ICPs showed more than 80% sensitivity and 75% specificity. Both statistics were more than 80% when the sampling fractions were more than 25% in ICP.

**Table 1:** Sensitivity and specificity of 100 replicates of simulations in detecting high-risk Medical Officer of Health areas with classical and inhibitory geostatistical design with close pairs (ICP) design

| Sample size | Sensitivity (95% Interval) | Specificity (95% Interval) |
| --- | --- | --- |
| Classical design |  |  |
| 563 (50%) | 85.5 (83.4 – 87.6) | 86.5 (84.2 – 88.7) |
| 372 (33%) | 83.2 (80.6 – 85.8) | 87.8 (85.3 – 90.3) |
| 282 (25%) | 80.7 (77.9 – 83.5) | 85.2 (82.5 – 88.0) |
| 225 (20%) | 80.7 (77.4 – 84.1) | 85.4 (83.0 – 87.7) |
| 169 (15%) | 78.9 (74.8 – 83.1) | 80.7 (77.8 – 83.6) |
| 113 (10%) | 73.5 (68.6 – 78.5) | 79.5 (75.8 – 83.1) |
| ICP design |  |  |
| 563 (50%) | 86.5 (84.8 – 88.3) | 81.3 (78.8 – 83.7) |
| 372 (33%) | 86.8 (84.9 – 88.6) | 80.9 (77.9 – 84.0) |
| 282 (25%) | 87.0 (85.1 – 88.9) | 80.9 (81.3 – 84.4) |
| 225 (20%) | 84.7 (82.1 – 87.2) | 81.7 (78.3 – 85.0) |
| 169 (15%) | 83.2 (80.3 – 86.1) | 77.6 (74.1 – 81.1) |
| 113 (10%) | 83.6 (80.9 – 86.3) | 75.0 (71.2 – 78.8) |

### Distance

Figure 5 plots the average percentage change in travel distance from reducing the sample size to different fractions of the original for both classical survey and ICP designs using the travel distance from the NSS as the base. Reducing the number of clusters leads to a substantial reduction in average travelling distance. For instance, reducing the number of clusters to 25% of the original sample resulted in a 44% reduction in average travel distance. The reductions in travelling time with the ICP design were slightly smaller than the reductions with classical survey design as ICP imposes a minimum distance between clusters to ensure spatial representativeness. However, in all cases the 95% confidence limits contained one, indicating that the ICP designs did not increase travel distance by comparison with classical designs.

**Figure 5:**
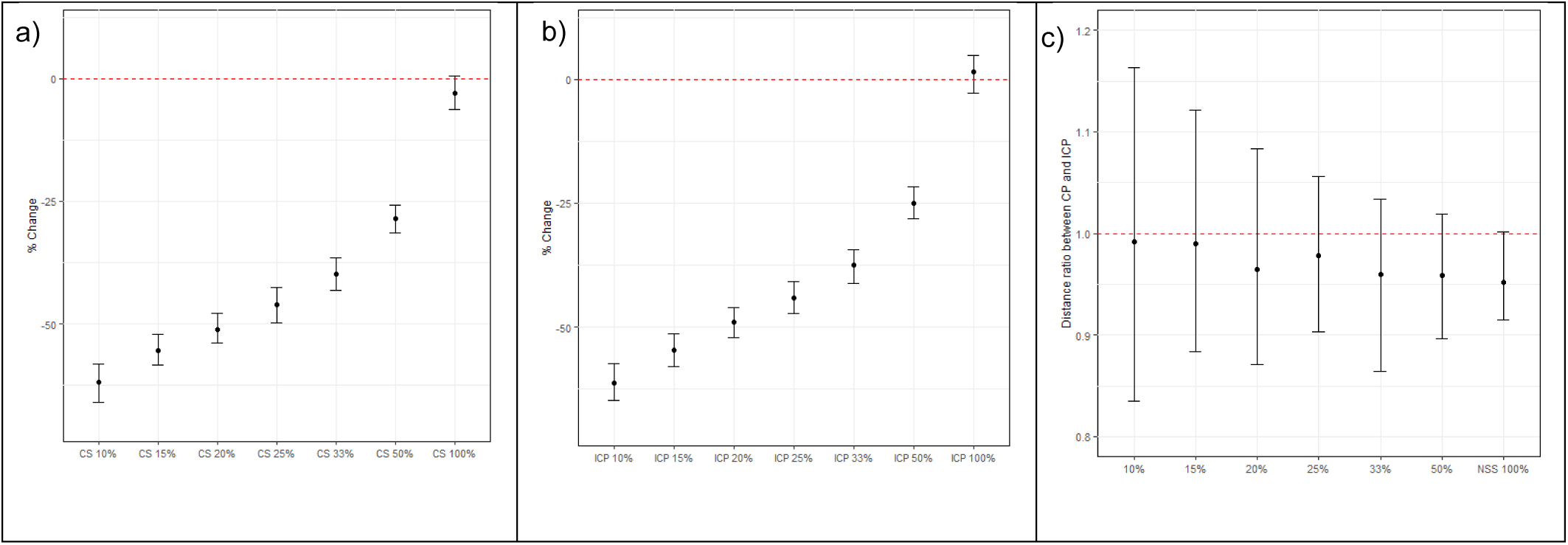
Change in mean travel distance. The average change in mean travel distance of 100 replications of simulations a) classical design, b) inhibitory close pairs (ICP) design and c) ratio between travel distance in classical and ICP designs at 10%, 15%, 20%, 25%, 33%, 50% and 100% of the original sample size in the National Snakebite Survey (NSS).

### Time taken to complete the survey

Both Classical design and ICP design showed a reduction in time duration to complete the survey at each fraction of the NSS sample size. The 95% confident limits were very narrow due to the high number of participants without bites. The lower 95% confident limits of the ratio between time taken to complete the survey in classical and ICP designs were greater than one at sampling fractions less than 50%, indicating that the ICP design can save time by comparison with the classical design.

**Figure 6:**
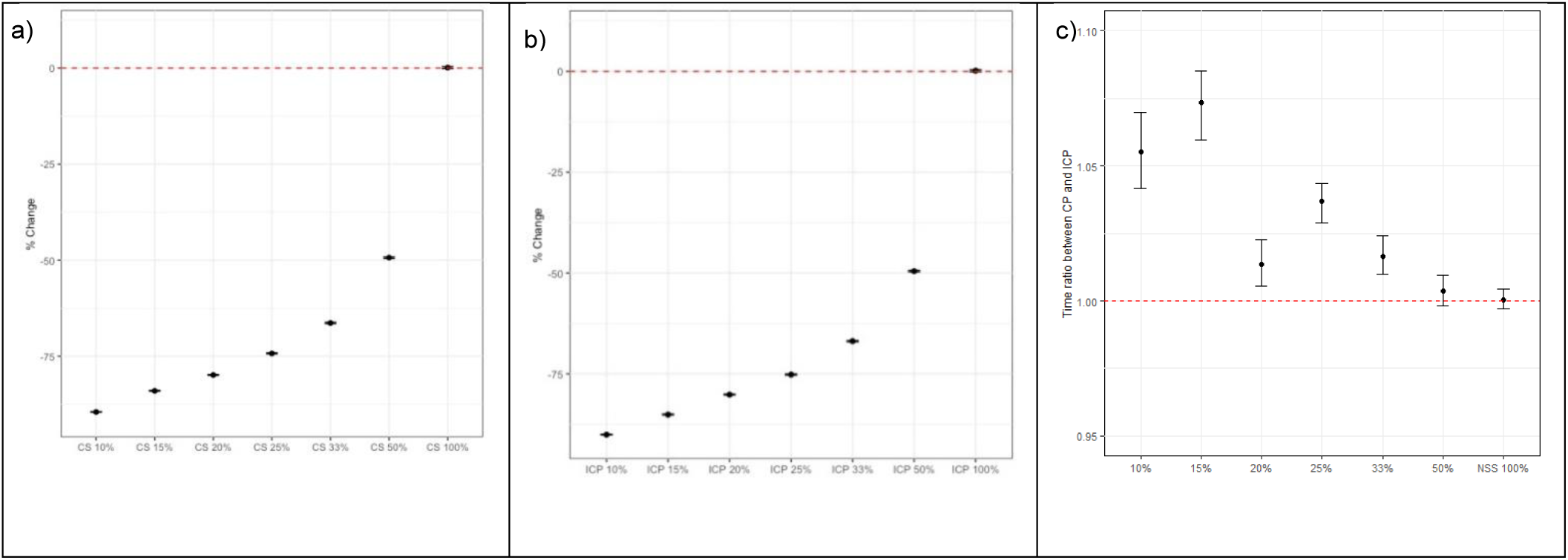
Change in mean survey time. The average change in mean time taken to complete the survey of 100 replications of simulations a) classical design, b) inhibitory close pairs (ICP) and c) ratio between time taken to complete the survey in classical and ICP designs fraction of the NSS sample size design corresponding to 10%, 15%, 20%, 25%, 33%, 50% and 100% of the original sample size in the National Snakebite Survey (NSS).

## Discussion

In this study, we compared the classical survey design with ICP in order to assess the utility of the ICP design in estimating snakebite envenoming incidence. Our results showed, firstly, that a similar predictive efficiency can be obtained at lower sample size with both the classical survey and ICP designs. This is a consequence of the spatial correlation in the incidence surface, whereby, *provided that the data are analysed using geostatistical methods*, the observed incidence of snakebite at any one location is predictive of the underlying risk in neighbouring locations. The classical survey design could yield almost the same predictive efficiency at *one third* of the original sample size, indicating the NSS has oversampled the country’s population. By comparison, the ICP design was able to produce the same results at *one fourth* of the original sample size of the NSS, highlighting an advantage of adopting ICP over classical survey design in estimating envenoming bite incidence in the country. Geostatistical methods, including ICP, are designed for efficient disease mapping over a region of interest and so comparatively smaller sample sizes are required compared to non-spatial, conventional survey sampling methods ^6^. Further, ICP showed a lower predictive variance than classical designs at all the sampling fractions, indicating ICP designs were able to produce more reliable predictions than conventional survey sampling methods. Therefore, resource requirements can be reduced by adopting ICP and geostatistical analysis in epidemiological surveys.

The smallest health care administrative divisions in Sri Lanka are the MOH areas. The NSS estimated the national envenoming bite incidence in Sri Lanka as 151 per 100 000 people. We classified the MOH areas as high risk and low risk based on the national envenoming bite incidence (i.e. gold standard). This was done to assess the sensitivity and specificity of ICP design in detecting high-risk MOH divisions. The ICP designs showed more than 80% sensitivity and specificity even at 25% of the original NSS sample size, showing their practical utility in snakebite research. Therefore, geostatistical methods, including ICP, can be considered a viable alternative for epidemiological surveys, particularly in low resource settings.

According to the NSS, snakebites show a substantial spatial correlation in Sri Lanka ^3^. Therefore, samples obtained from close locations can be expected to provide essentially the same information. Sampling methods based on random sampling can easily select sampling units close to each other which could be disadvantageous in these circumstances ^11^. Further, conventional survey sampling methods may be inefficient for predicting local risk, as the estimates can be associated with high standard errors, especially in areas with lower sampling efforts. The adoption of spatially regulated designs such as ICP has been proposed to lower the mean square prediction errors and reduce uncertainty in predictions ^7^. According to previously published work on snakebite in Sri Lanka, the spatial correlation structure had a non-negligible unexplained non-spatial variability due to a combination of spatial variation on a scale less than the minimum distance between two sampled locations and any measurement error arising from information bias. The inclusion of close pairs in the ICP design enables estimation of the full covariance structure, including both spatial and non-spatial random effects.

In ICP design, primary sampling units maintain a minimum distance between two units, which implies that a reduction in travel time to complete the survey does not necessarily follow a reduction in sample size. Therefore, we compared the total travel distance in ICP designs and the classical survey designs at different sampling fractions of original sample size of NSS survey. Our analysis showed ICP did not increase the travel distance compared to classical survey designs. However, since the required sample size is less in ICP, this will, in turn, reduce the travel-related costs incurred to complete the survey. Similarly, we evaluated the time taken to complete the survey, and ICP showed a lower time to complete the survey by comparison with conventional survey sampling methods at sampling fractions. Also, time taken to complete the survey had also reduced as a consequence of the smaller required sample size.

In conclusion, our study showed that snakebite envenoming incidence can be estimated with similar precision to classical survey designs such as multi-stage cluster sampling by adopting an ICP design with smaller sample size and travel time. This would substantially save resources and time taken to conduct surveys. ICP should be considered for epidemiological studies in low resource settings. However, there are challenges to adopting geostatistical methods. Firstly, an ICP design strictly requires prior geolocations of all units in the sampling frame. In the absence of these, a pragmatic solution is the following. A provisional ICP design is constructed in which every point in the design region is considered as a potential sampling location. The field team is then asked to identify the available sampling location closest to each point in the provisional design. A second limitation is that expertise in geostatistical analysis methods is needed to get the maximum benefit from georeferenced data. This highlights the importance of knowledge-sharing across the scientific community, especially in low-resource settings ^11^.

## Data Availability

All data produced in the present study are available upon reasonable request to the authors

## Acknowledgements

The authors wish to acknowledge the research team who designed and undertook the National Snakebite Survey.

## Ethics approval

Ethical approval for the National Snakebite Survey was obtained from the Ethics Review Committee of the Faculty of Medicine, University of Kelaniya. All interviews were conducted after obtaining informed written consent. Permission for conducting the survey was obtained from District- and Divisional-level public administrators. This study used only the secondary data.

## Author contributions

DSE, TDS, AK, HJDS and PJD designed the Study. DSE and TDS designed the analytical strategy, conducted the analysis and interpret the results. DSE drafted the manuscript and TDS, AK, HJDS and PJD critically revised the article. All approved the final version.

## Data Availability

The data underlying this article will be shared on reasonable request to the corresponding author.

## Funding section

### Funding

None

